# Comparative Prediction of Psychotic and Mood Disorders with Multi-Model Machine Learning

**DOI:** 10.1101/2025.08.13.25333573

**Authors:** Mercy O. Oyebode, Racheal I Rieninwa, Samuel Okodeh, Isioma Aniukwu, Ephraim O. Nwoye

## Abstract

**Introduction:** Recently, there has been a surge in the number of mental health cases including paranoid schizophrenia (psychosis) and depression (mood disorder).

**Objectives:** This study conducted a comparative prediction of psychotic and mood disorders using multi-model machine learning (MLs), mainly: Logistic Regression (LR), Support Vector Classification (SVC), Random Forest (RF), and Extreme Gradient Boost (XGBoost). Methods: The study used a clinical datasets of 318 patients diagnosed with Psychotic and Mood related disorders. We created a mid-category of Psychotic-Mood disorders for properly classifying patients having shared symptoms with Psychotic and Mood disorders. The Column Transformer preprocessor was used for modeling. The categorical columns were encoded using One-Hot-Encoder and Imputed using Simple Imputer with most-frequent as the strategy. The numerical columns were also encoded using Standard Scaler and imputed using Simple Imputer with mean as the strategy.

**Results:** Our results showed a consistent performance hierarchy in the confusion matrices (RF > XGBoost ≈ SVC > LR) for pure conditions, contrasted with the reversed pattern for mixed conditions, computationally validating long-standing clinical debates about psychiatric nosology and supports dimensional rather than categorical approaches, particularly highlighting that algorithmic complexity does not necessarily improve classification of inherently unstable diagnostic categories.

**Conclusion:** We found that while conventional diagnostic criteria seeks to clearly differentiate psychiatric conditions, our computational evidence supports that psychotic-mood disorders often fall into a spectrum of conditions and that psychiatric comorbidity patterns can be detected using machine learning models.

## 1. Introduction

In recent times, there has been a rise in the number of mental health diseases such as paranoid schizophrenia (psychosis) and depression (mood disorder). According to the National Institute of Mental Health, Psychosis is a mental health disorder characterized by an individual disconnected from reality. The causes could be attributed to genetic risks, health conditions, medication or drug use. In Nigeria, there are over 100 thousand cases of psychoses per year. For instance, the stress-vulnerability model reveals that there are various causative factors of schizophrenia, including social, psychological and biological (Quaedflieg et al., 2013, as cited by Nwoye et al., 2024). Typical symptoms of psychosis and mood disorders include schizophrenia, bipolar disorders, abnormal behaviour, etc.

In a quest to address the increasing mental health conditions challenges, advanced computing techniques such as Artificial Intelligence (AI) and Machine Learning (ML) are emerging as viable potential solutions through implementing data-driven paradigms (Squires et al., 2023). Machine Learning, the study and development of statistical algorithms that can learn from data and be used for the generalisation of unseen data (predictive analytics), along with deep learning and artificial neural networks, stands a good chance of contributing positively to improving an individual’s mental health conditions. Adopting predictive analytics in healthcare holds many benefits and raises many individuals’ hope of recovering from mental illness through early detection, correct care and proper treatment (Chung and Teo, 2022).

Today, there exists a good number of ML models. Madububambachu et al. (2024) outlined in their review how various algorithms (ML models), including Decision Tree, Neural Network, Support Vector Machine, Naive Bayes, and logistic regression algorithms, were used to group students having different mental health problems. It was revealed that distinct models performed optimally for specific health concerns. For instance, Zaccheus, Fidelis, and Nwoye (2021) developed a neural network model for schizophrenia diagnosis. The model achieved a 90% accuracy which was high enough to develop a software with the potential to perform schizophrenia diagnosis. Such software can help in providing individuals-specific treatment plans using predictive features to enhance patient outcomes. Agnihotri and Prasad (2024) in a previous work also demonstrated that a hybrid LRSVM ML model was good at predicting bipolar disorder by combining the strengths of LR and SVM to ensure accurate and reliable predictions. Ndikumana et al. (2025) further conducted research in predicting Rwanda’s youth mental health using SVM, LR, RF and XGBoost models.

This research work focuses on conducting comparative prediction of psychotic and mood disorders using multi-model MLs, namely, Logistic Regression (LR), Support Vector Classification (SVC), Random Forest (RF), and Extreme Gradient Boost (XGBoost). While previous works have shown that different models perform better with specific diagnostic groups, this research attempts to capture the diagnostic relationships between psychotic and mood disorders. In real-world scenarios, psychiatric conditions often demonstrate overarching symptoms and comorbidities. This approach was more practical in this research due to the limited size of the available clinical dataset. The result from Ndikumana et al. (2025) showed that the Random Forest model outperformed SVM, LR and XGBoost. Confusion matrix and Feature importance were used as the metric scores to validate the accuracy of their models. However, as a result of the nature of the dataset, their analyses were limited to biological and social factors. Other limitation factors include dataset class imbalance. To address this challenge in this study, the Synthetic Minority Oversampling Technique (SMOTE) was used. SMOTE is a technique that performs interpolation between minority classes and their K-nearest neighbours. This is used to generate synthetic data patterns (Elreedy et al., 2024). This had a significant improvement in the accuracy of the models in this study. Wijaya and Rachmat (2024) also utilised the SMOTE technique in their research. Their findings revealed that Logistic Regression, which had the best performance, had an improvement in F1 Score accuracy from 0.8282 to 0.8337 when SMOTE was used.

For this study, thematic analysis was applied in grouping the dataset. Thematic analysis involves the identification and reporting of patterns in a dataset (Naeem et al., 2023). By learning from patterns in the complete data, AI models are trained to impute missing categorical data. The data is analysed based on smaller clinically related groups or blocks, instead of imputing all the missing values at a go. For example, Do et al. (2023) in their study applied the Blockwise Principal Component Analysis (PCA) framework to separate single tone blocks and separately imputed each block. This method reduces dimensionality and improves imputation accuracy and speed. Similarly, Héctor A. et al. (2024) applied an EM-based block-by-block imputation approach to ADNI’s multi-source clinical data, which leads to improved signal recovery and completeness in the final dataset.

## 2. Methodology

This study focused mainly on Psychotic and Mood disorders. However, it also incorporated datasets grouped in Psychotic-Mood disorders categories due to their shared symptoms with Psychotic and Mood disorders. According to American Psychiatric Association (2022), schizoaffective disorder is characterised by a combined occurrence of mood episodes and the active phase symptoms of schizophrenia. Similarly, Parker (2014) stated how bipolar disorders have been re-categorised into the psychotic-mood disorders classification. This is because it has been recognised that bipolar disorders form a spectrum between the two diagnostic classes rather than two distinct classes in terms of symptoms overlap, family history and genetics.

Other diagnosis categories were not included since this research work focuses on psychotic disorders, mood disorders and psychotic-mood disorders. These categories are commonly grouped under broader diagnostic groupings in International Classification of Diseases, ICD-11, for improvement and clarity in analytical and machine learning modelling (Gaebel W. et al., 2020).

This study used a monocentric clinical dataset acquired from the Federal Neuropsychiatric Hospital, Lagos, Nigeria, with approval from the ethical committee of the hospital. The data contained a total of 664 health records containing various psychiatric diagnostic conditions from year 2013 to year 2019.

### 2.1 Data cleaning

The data features include: Diagnosis, Sex, Occupation, Marital status, Duration of the current episode (likely in days, weeks, or months), Past psychiatric history, Past medical history, among others. Out of the 38 features, the Thought Possession column was dropped as it contained more than 50% missing values. The class feature was dropped as it has a relationship with the Diagnostic feature, which could contribute to data leakage in the models.

The Diagnosis feature Column, which is the target variable, has about 50 categories of diagnosis. These were afterwards structured into 3 major groups: Psychotic Disorders (71%), Mood Disorders (18%), and Psychotic-Mood Disorders (11%). Paranoid Schiz, Hebephrenic Schiz and Simple Schiz cover 82% of the Psychotic Disorder class. The Mood class was characterised mainly by Depression, while the Psychotic-Mood Disorder class included Bipolar Affective Disorder, Schizoaffective Disorder and Acute Psychosis.

Thematic analysis was utilised to group the features into four broad categories, namely: Mental Status Examination (MSE), Psychiatric & Personal History, Investigative / Clinical Interview, and Diagnosis & Functionality Group, as shown in Table 1.

**Table 1.** The Blockwise Imputation Groups.

| Thematic Block Group | Indicators | Applicable Features |
| --- | --- | --- |
| Mental Status Examination | assess the patient's current mental state, such as mood, thought process, perception, memory, attention | MOOD, AFFECT, INSIGHT, TH_FORM, TH_CONTENT, TH_STRM, PERCEP, SPEECH, MEM_LT, MEM_ST, MEM_IR, CONC, ORIENT, ATTEN, JUDGMT \| |
| Psychiatric & Personal History | Captures the patient's clinical, social, and personal history | P_SEX_HX, P_MED_HX, PREMOBD_HX, P_PSY_HX, P_SOC_HX, FOR_HX, FAM_P_HX, EEG |
| Investigative / Clinical Interview | These are insights or scores from clinical interviews or structured diagnostic assessments. | TH_POSS, INT_PROV, INT_CAL, INT_S_A_D, INT_GFK |
| Diagnosis & Functionality | These reflect final outcomes, functioning, and high-level status markers | DIAGN, MSE, DUR_EPIS, MAR_STA, OCCUP |

Based on this grouping, an AI-driven imputation approach was adopted in handling the missing values. Mode imputation for columns with low missing-ness (<10%), Random Forest Classifier for moderate missing-ness (10%–40%) and more than 40% were dropped from the data. After these blocks were extracted for cleaning and imputation, they were combined and added to the remaining data.

### 2.2 Preprocessing

The Column Transformer preprocessor was used for modeling. The categorical columns were encoded using One-Hot-Encoder and Imputed using Simple Imputer with most-frequent as the strategy. The numerical columns were also encoded using Standard Scaler and imputed using Simple Imputer with mean as the strategy.

### 2.3 Setup of computational experiment

To achieve a comparative prediction of psychotic and mood disorders diagnosis, the 4 ML models were trained and tested with 318 data points using 36 features, varying several train/test ratios (90/10, 80/20, 70/30, and 60/40) to obtain the best configuration. The 70/30 configuration produced the best accuracy with the least degree of over-fitting. The models were initially configured with custom parameters to address dataset-specific challenges and thereafter hypertuned to achieve better results. The ML models were trained using Python on Anaconda IDE with inbuilt libraries such as Pandas, Scikit-learn (sklearn), Imbalanced-learn (imblearn), XGBoost, Numpy and Matplotlib, etc. All experimentation was carried out on a Lenovo ThinkPad having an Intel(R) Core(TM) i5-6200U CPU @ 2.30GHz 2.40 GHz processor and 256GB SSD.

## 3. Results

The Machine Learning multi-models used are Logistic Regression (LR), Support Vector Classification (SVC), Random Forest (RF), and Extreme Gradient Boost (XGB) models. The models were trained and modelled using default SMOTE and Class weights. The models were then evaluated using Cross-validation F1 Score. The model result below demonstrated overfitting and class biased. The F1-macro scores obtained for the various models before hyper-tuning was in the range of 0.33 to 0.49. The result is shown in Table 2.

**Table 2.**
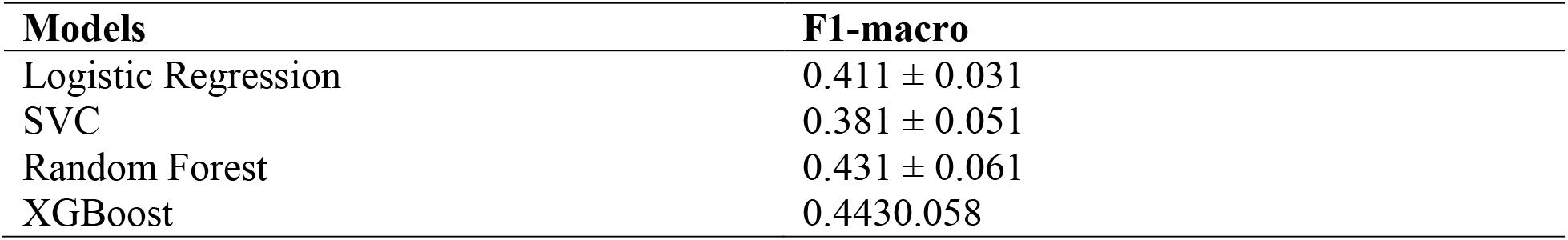
Cross-validation F1 scores for the four models before tuning.

### 3.1 Hyperparameter tuning

In order to improve the accuracy of the models above, individual pipelines were built for each of the models instead of building generalized pipelines. Each of the models were then reconfigured using model-based pipeline and hyperparameter tuning using grid search. The results showed better model accuracy. The best parameters for Logistic Regression were: C = 1, penalty = l2, solver = lbfgs. The SVC was optimized with the following parameters: C = 100, gamma = 0.01, kernel = rbf. Best parameters for Random Forest were: max-depth = 20, min_samples_split = 2, n_estimators=200. XGBoost optimized with: colsample_bytree 1.0, learning rate = 0.1, min_child_weight = 4, n_estimators = 200, and subsample = 0.8.

The F1 Macro results and the Macro AUC results when the models parameters were tuned are shown Table 3. AUC is the area under a receiver operating characteristic (ROC) curve.

**Table 3.** Cross-validation F1 and AUC scores for the four models after tuning.

| S/N | Model | F1 Macro | Macro AUC |
| --- | --- | --- | --- |
| 0 | Logistic Regression | 0.6220 | 0.7583 |
| 1 | SVC | **0.6574** | 0.8103 |
| 2 | Random Forest | 0.6407 | **0.8652** |
| 3 | XGBoost | 0.6161 | 0.8315 |

The outcome of the comparative prediction showed that SVC had the highest classification accuracy with a F1 Macro Score of 0.6574. However, Random Forest had best discriminative ability with an AUC Macro Score of 0.8652. A mid-range F1 scores of 0.61 to 0.66 were observed across all the models, reflecting inherent diagnostic complexity in psychiatric grouping, especially when class imbalance is present. The AUC score range of 0.76 to 0.87 indicates higher potential for clinical decision support applications such as computer aided diagnostic assessments. To further visualize how the models behaved, ROC/AUC curves were generated and shown in Figure 1 – 4

**Figure 1.**
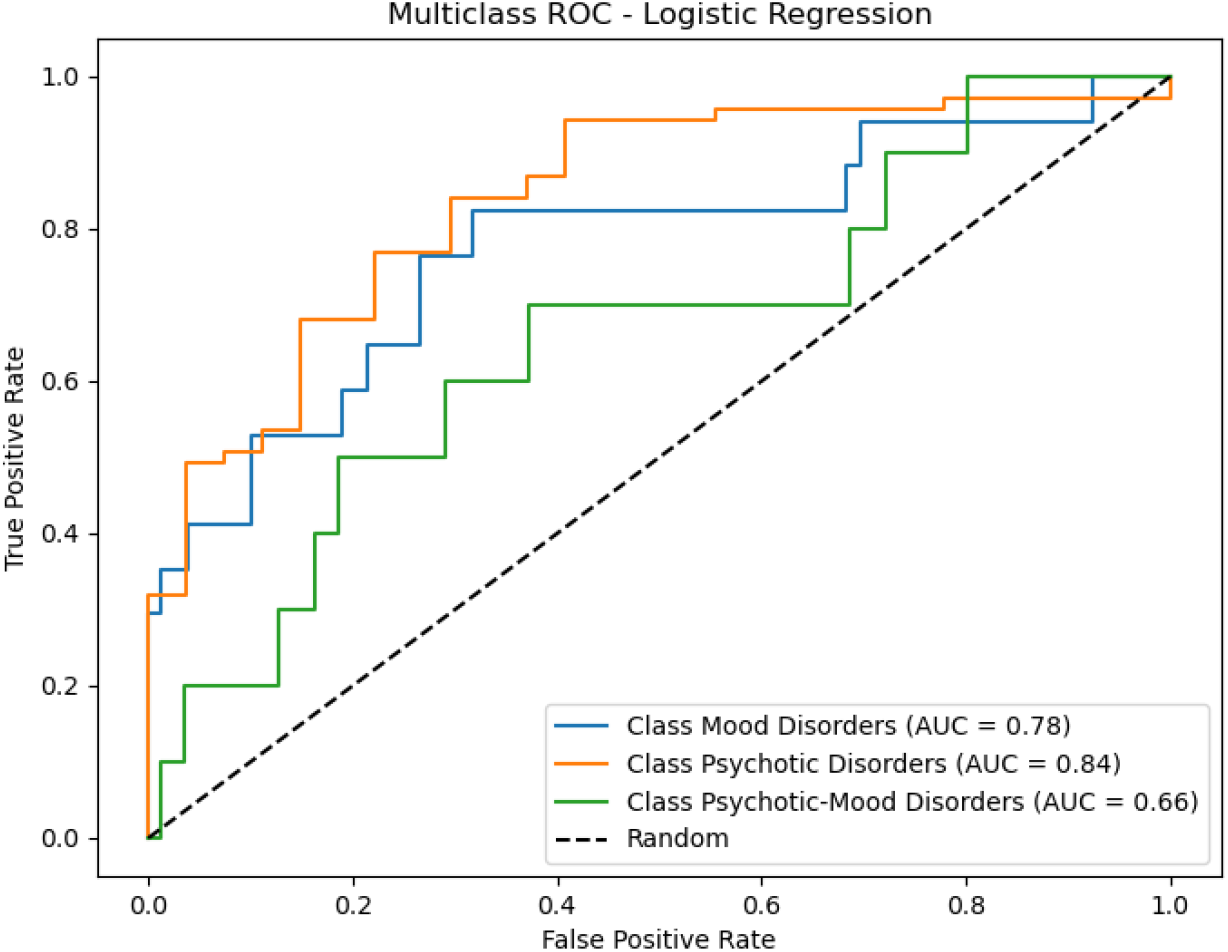
Area under the curve (AUC) for Logistic Regression model

**Figure 2.**
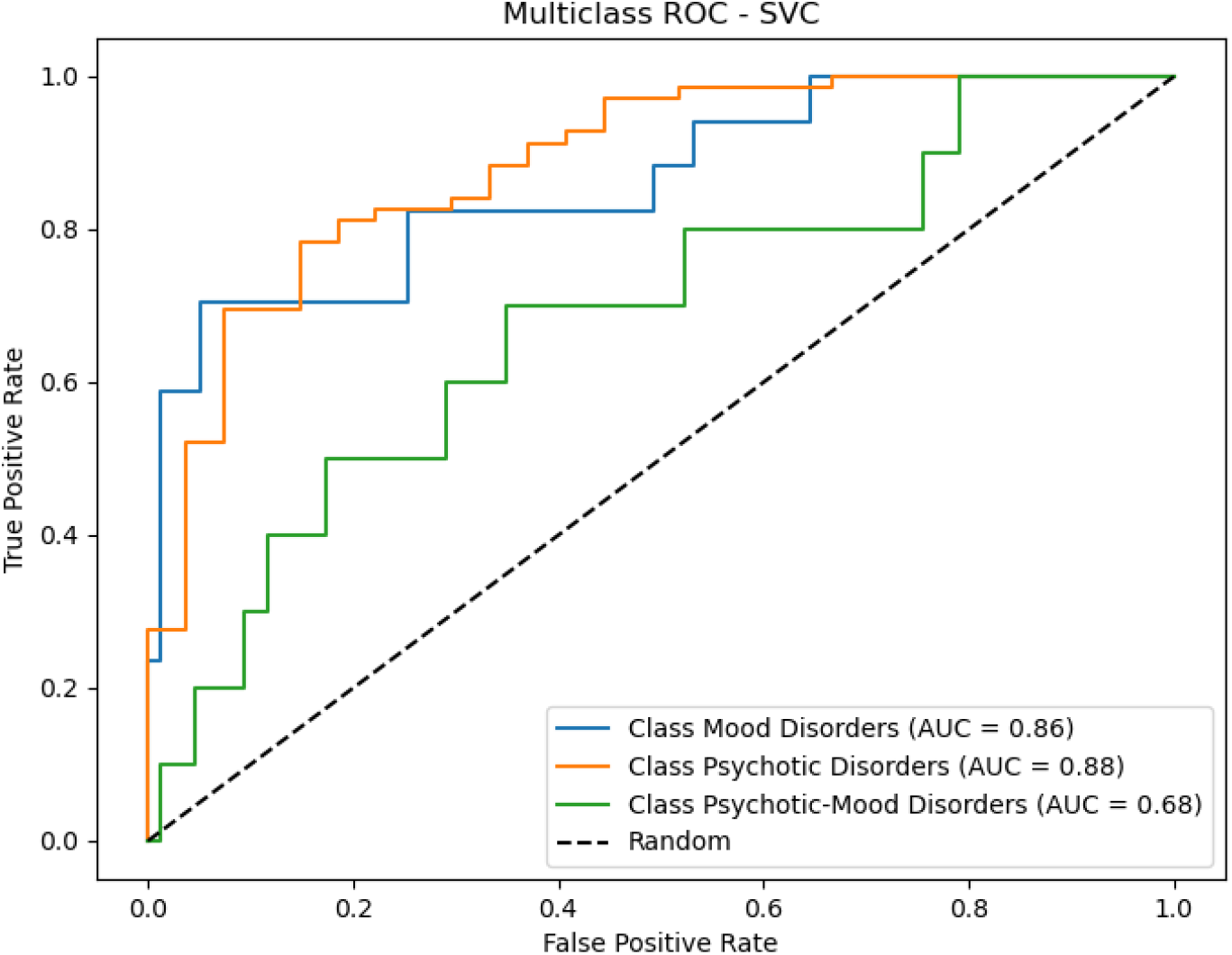
Area under the curve (AUC) for SVC model.

**Figure 3.**
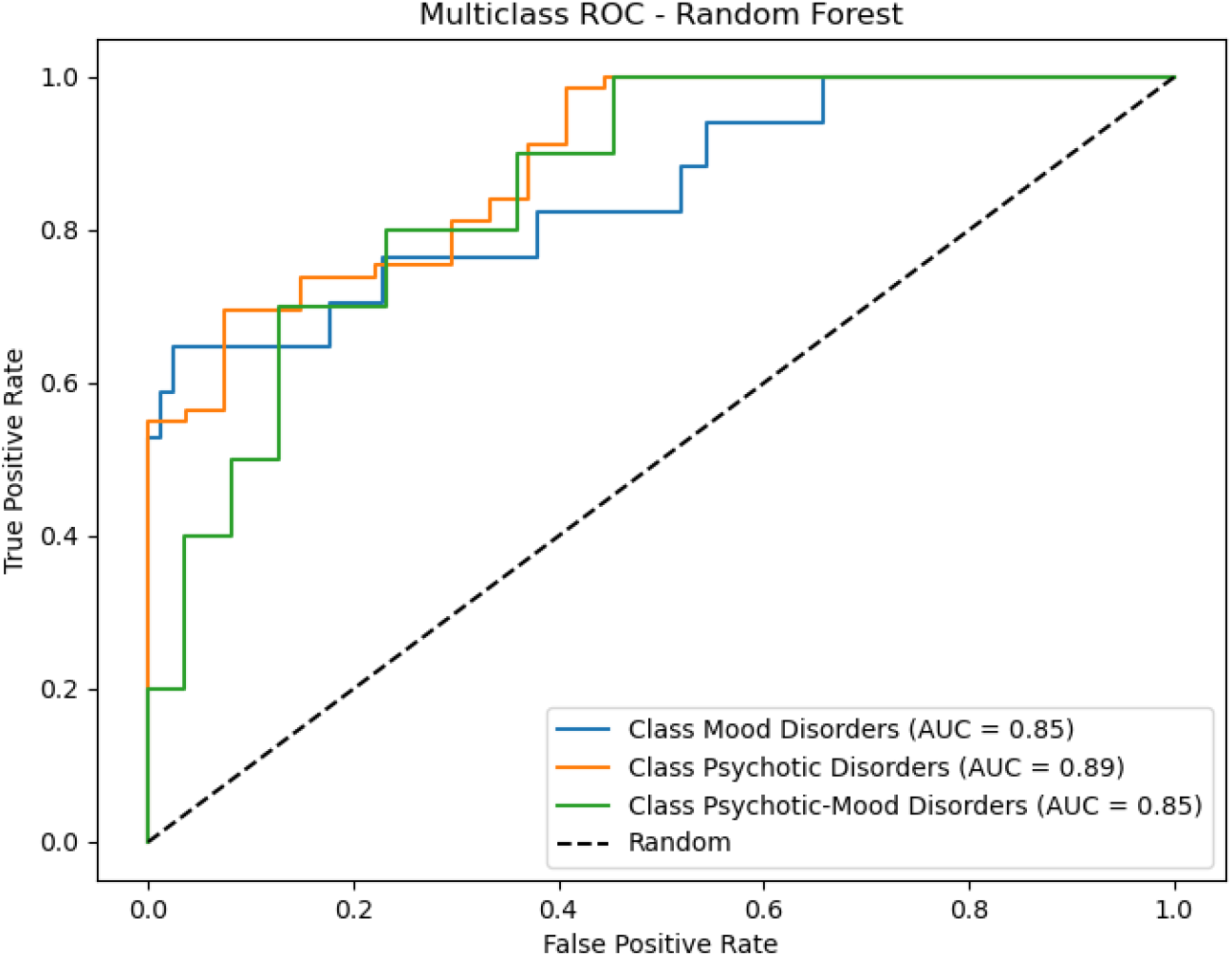
Area under the curve (AUC) for Random Forest model

The ROC Curve plots for the four models showed that the models could perform better when properly configured and the hyper-parameters are further tuned. The dashed diagonal line showed the minimum ROC Score of 50%. It implied the minimum score a model can get. The results are shown in Table 4 for better comprehension.

**Table 4.** Area under curve (AUC) for the four models.

| CLASS | Logistic Regression (AUC) | SVC (AUC) | Random Forest (AUC) | XGBoost (AUC) | F1 Best Score |
| --- | --- | --- | --- | --- | --- |
| Mood Disorders | 0.78 | **0.86** | 0.85 | **0.86** | 0.86 |
| Psychotic Disorders | 0.84 | 0.88 | **0.89** | 0.88 | 0.89 |
| Psychotic-Mood Disorders | 0.66 | 0.68 | **0.85** | 0.75 | 0.85 |

The Overall Macro Average ROC curve plot for the four models is shown in Figure 5. The plot revealed that the Random Forest model performs with an accuracy score of 0.87 and XGBoost model with accuracy score of 0.83. Future works include improving the model accuracy so that psychotic and mood disorders could be accurately predicted.

**Figure 4.**
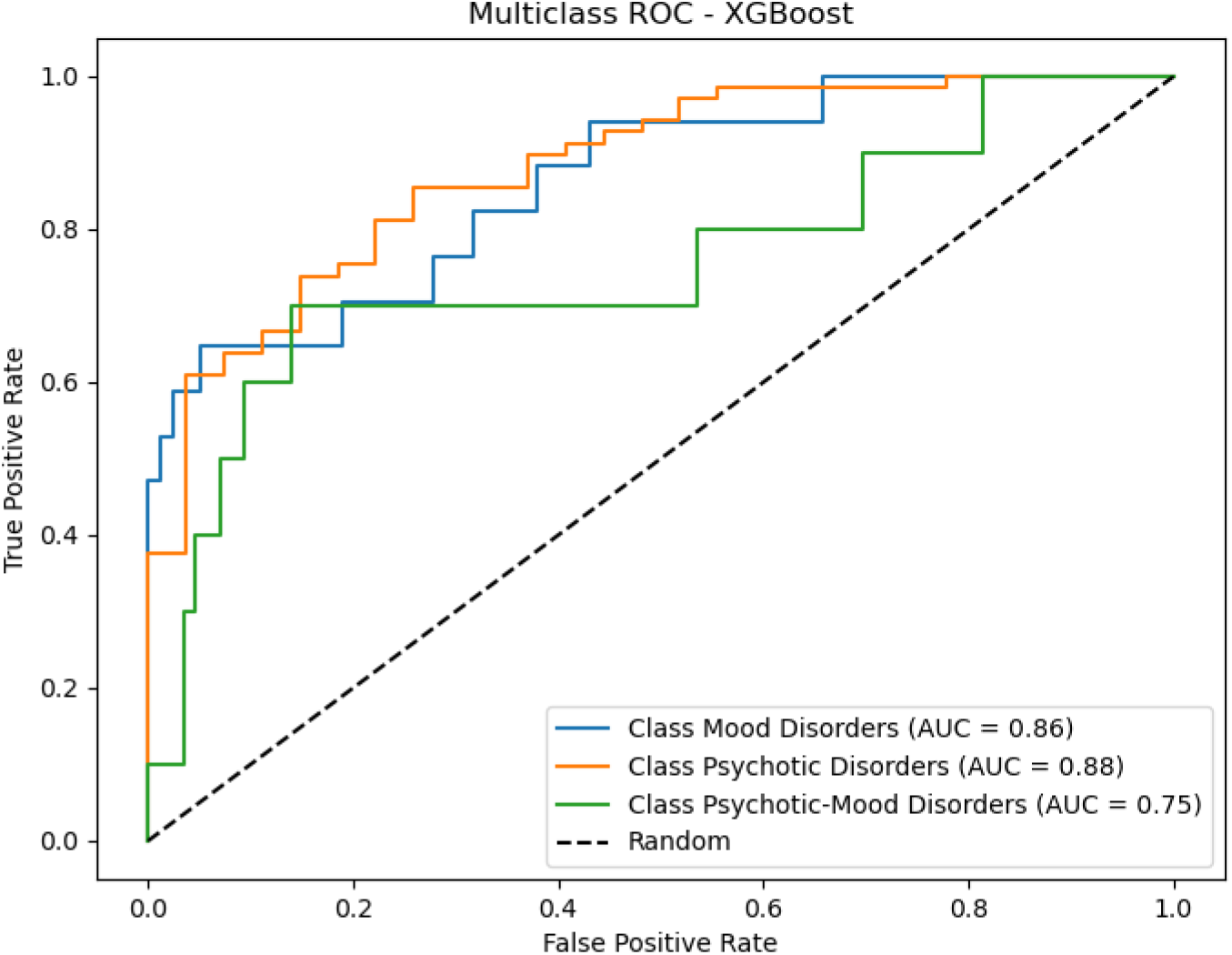
Area under the curve (AUC) for Extreme Gradient model

**Figure 5.**
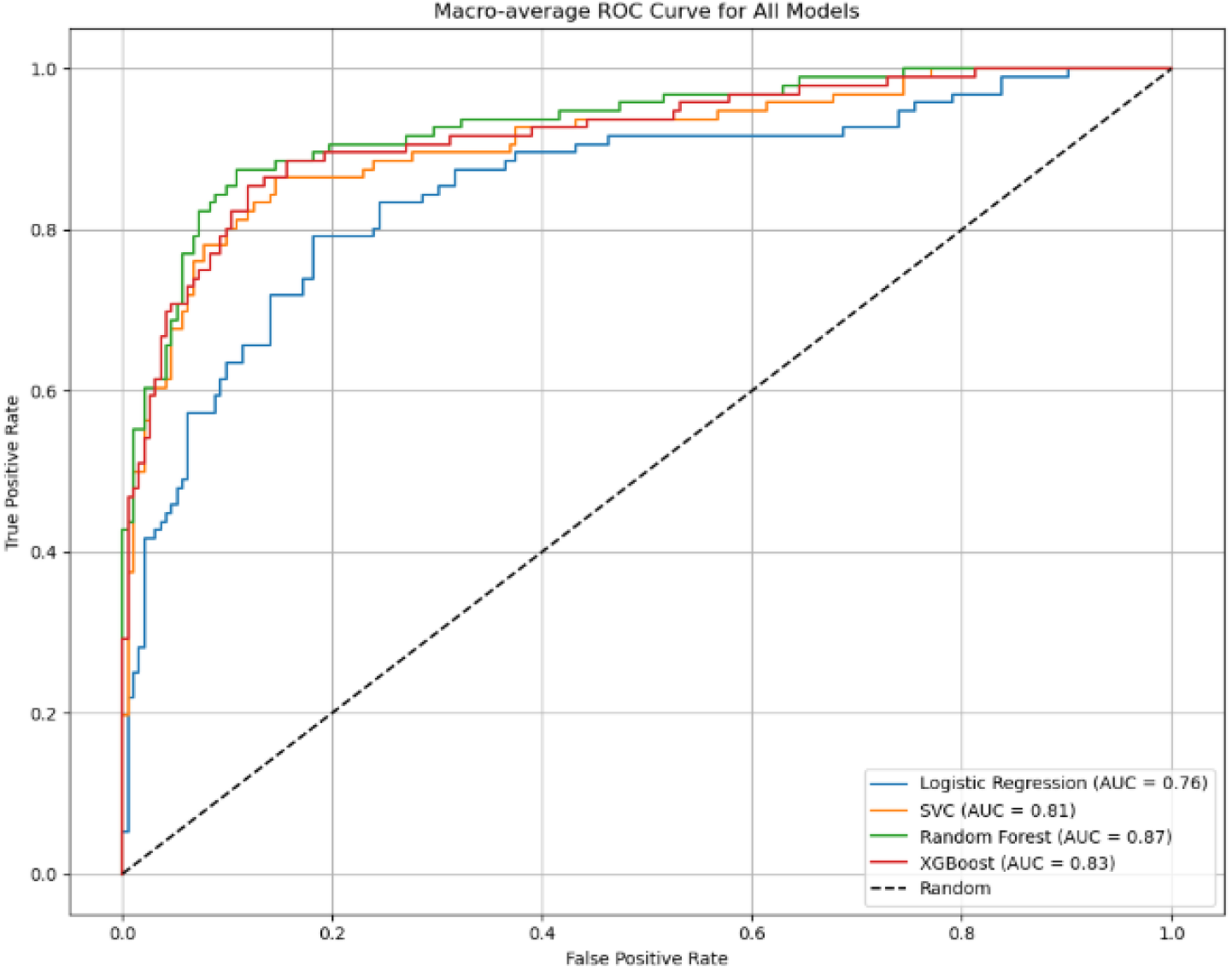
Area under curve for all models.

## 4. Discussions

In predicting psychotic and mood disorders using the 4 ML models, there was a need to validate the accuracy of these models. This was done through the aid of Confusion Matrix. The confusion matrices revealed consistent misclassification of Psychotic disorders, Mood disorders and psychotic-mood disorders in a minimal way. The confusion matrices for the four models are shown in Figure 6 – 9. Similarly, SHAP (Shapley Additive exPlanation) a based feature importance analysis was further used to investigate and mitigate the misclassification issues observed in the confusion matrices.

**Figure 6.**
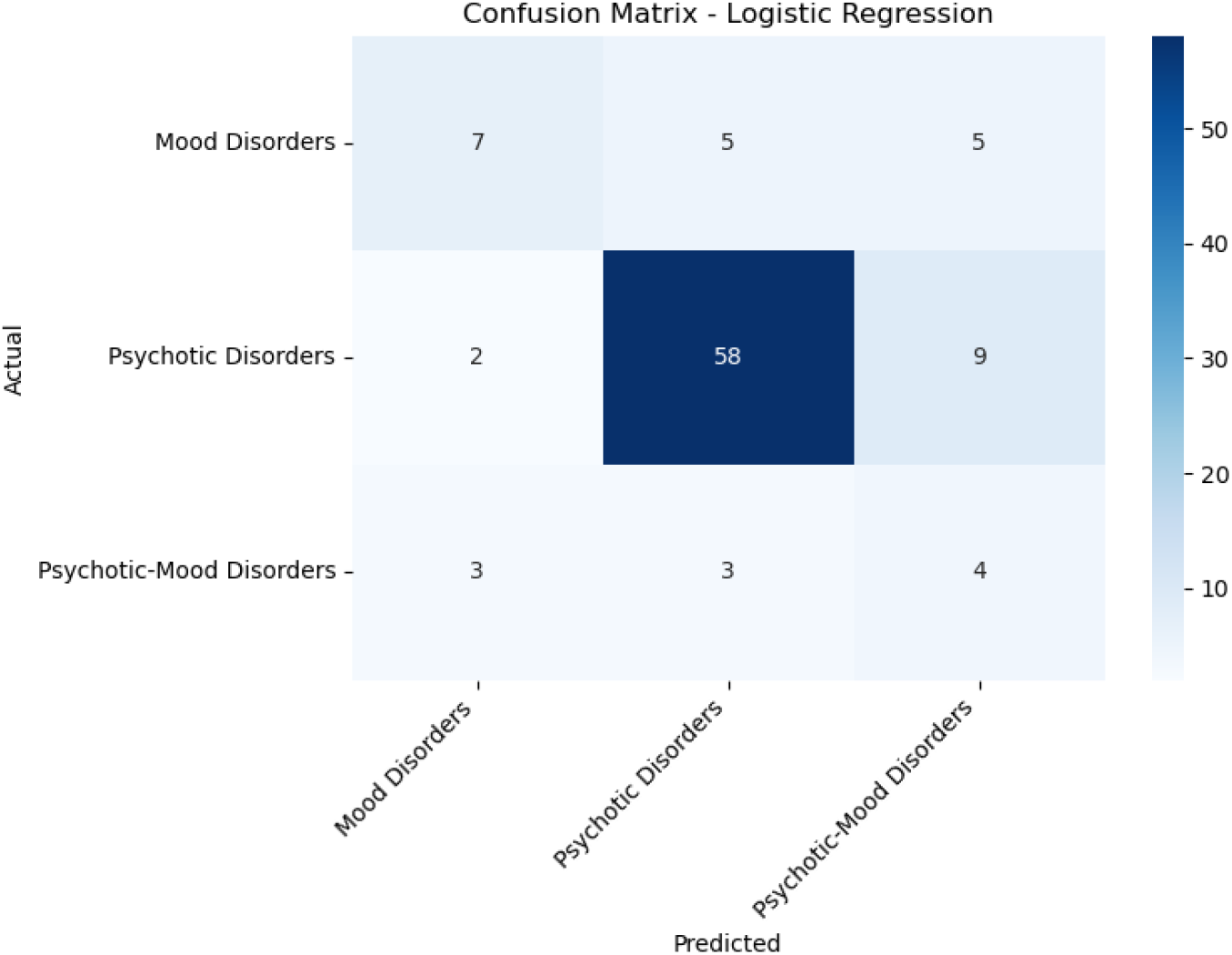
Confusion matrix for Logistic Regression model

### 4.1 Model comparison analysis across confusion matrices

The confusion matrix results revealed distinct performance patterns across all four models, with Random Forest demonstrating superior performance for pure Psychotic Disorders (98.6% recall) compared to XGBoost (92.8%), SVC (89.9%), and Logistic Regression (84.1%), reflecting how ensemble methods better capture the distinct and stable symptom profiles characteristic of schizophrenia spectrum disorders. For Mood Disorders, performance varied significantly across models: XGBoost achieved the highest recall (58.8%), followed by SVC (52.9%), Random Forest (64.7%), and Logistic Regression (41.2%), with all models showing 6-7 misclassifications as psychotic disorders, aligning with clinical reality where 18.5% of major depressive episodes present with psychotic features, creating consistent diagnostic boundary challenges across algorithmic approaches. Psychotic-Mood Disorders showed the most interesting pattern: while Random Forest and XGBoost both achieved only 20% recall, Logistic Regression surprisingly performed best with 40% recall, and SVC achieved 30%, suggesting that simpler linear models may paradoxically better capture mixed-condition patterns, though all models struggled with the diagnostic instability of conditions like schizoaffective disorder. The consistent performance hierarchy (Random Forest > XGBoost ≈ SVC > Logistic Regression) for pure conditions, contrasted with the reversed pattern for mixed conditions, computationally validates long-standing clinical debates about psychiatric nosology and supports dimensional rather than categorical approaches, particularly highlighting that algorithmic complexity does not necessarily improve classification of inherently unstable diagnostic categories.

### 4.2 Feature importance analysis using SHAP

SHAP: (Shapley Additive exPlanation) – SHAP is powerful as it helps to reveal how each feature impacts the models in order of importance, for instance Bee swarm plot. Y-Axis (Features): Features ranked by importance (top = most influential) and the X-Axis (SHAP Value) shows the Impact of that feature on the model’s output for a given prediction. Positive SHAP values drive prediction towards the class features while negative SHAP values drive prediction away from the class features. Each dot represents a single observation (patient) for that feature. The Red for High value feature, Blue for Low value feature and Purple color for Intermediate value feature.. Comparison between the Bee Swarm Plots of for all the models are shown in Figure 10 to 13. The plots revealed that features in the Mental Status Examination (MSE) Group, such as mood, insight, th_form, th_content, percep, conc, and judgmt show high values and contribute to prediction of the three classes, followed by the features in Demographic and Functionality Group such as sex, marital status, occupation etc. then to features in Psychiatric & Personal History such as premorbid_hx, p_med_hx and p_psy_hx.

**Figure 7.**
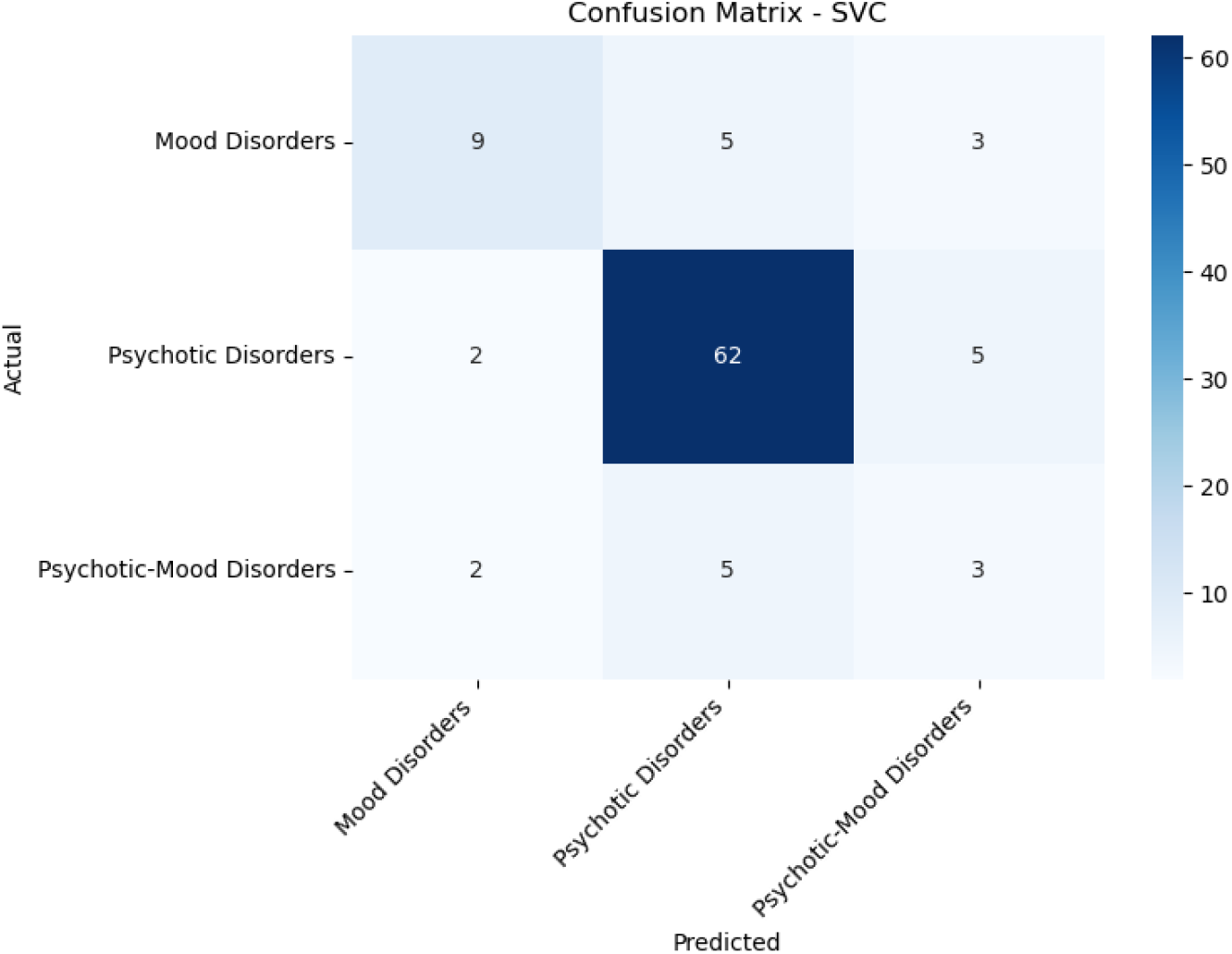
Confusion matrix for Support Vector Classification model

**Figure 8.**
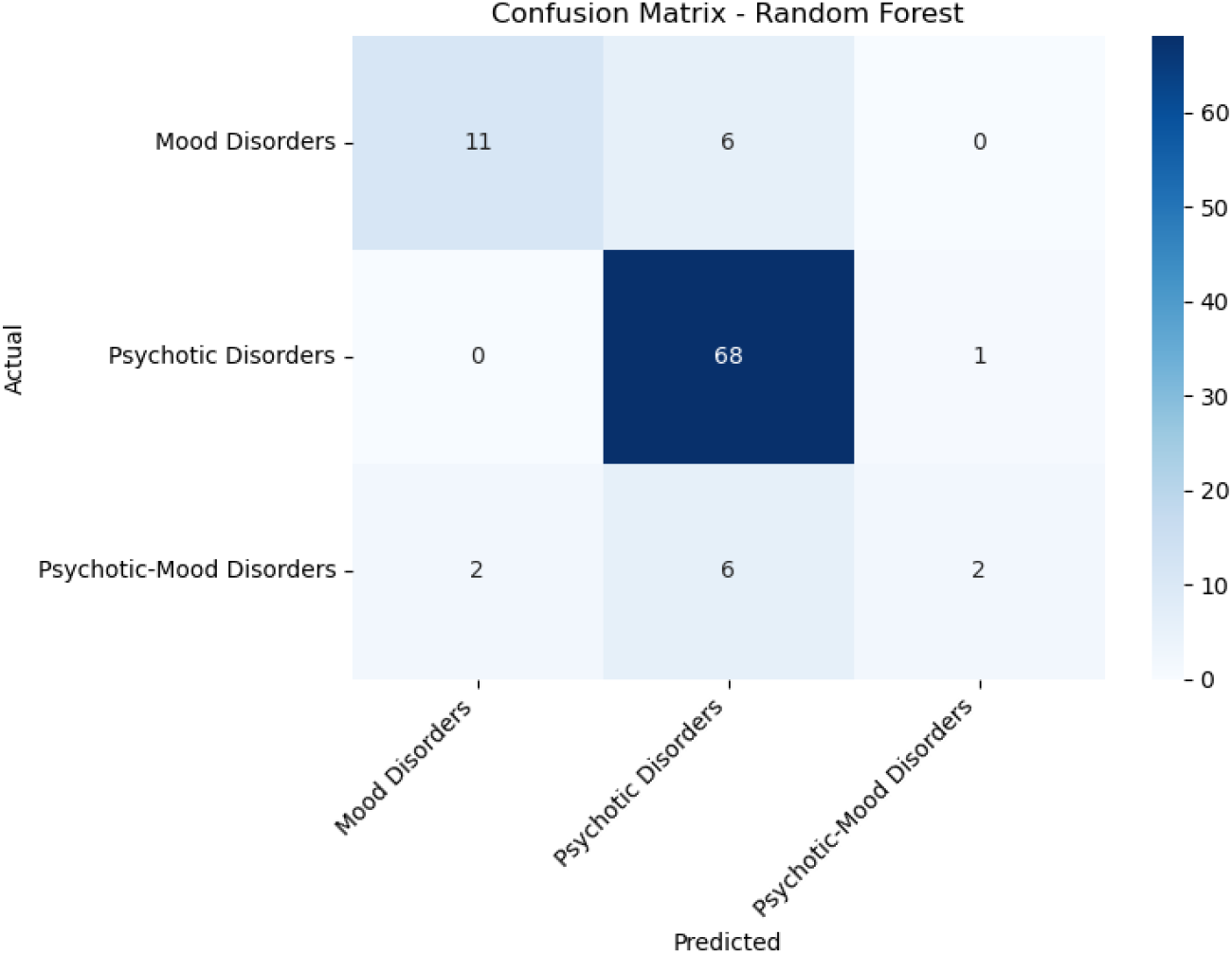
Confusion matrix for Random Forest model

**Figure 9.**
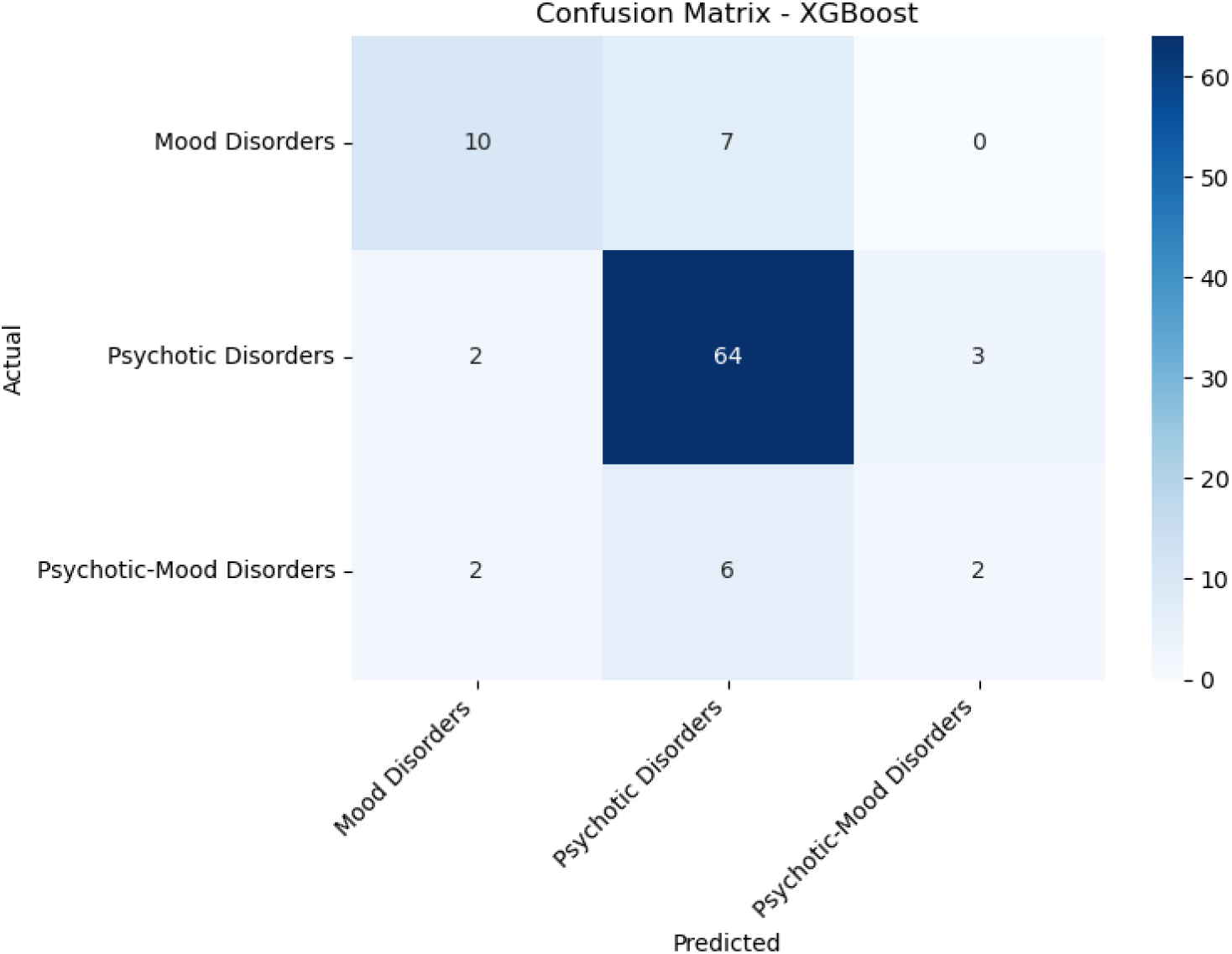
Confusion matrix for Extreme Gradient model

**Figure 10.**
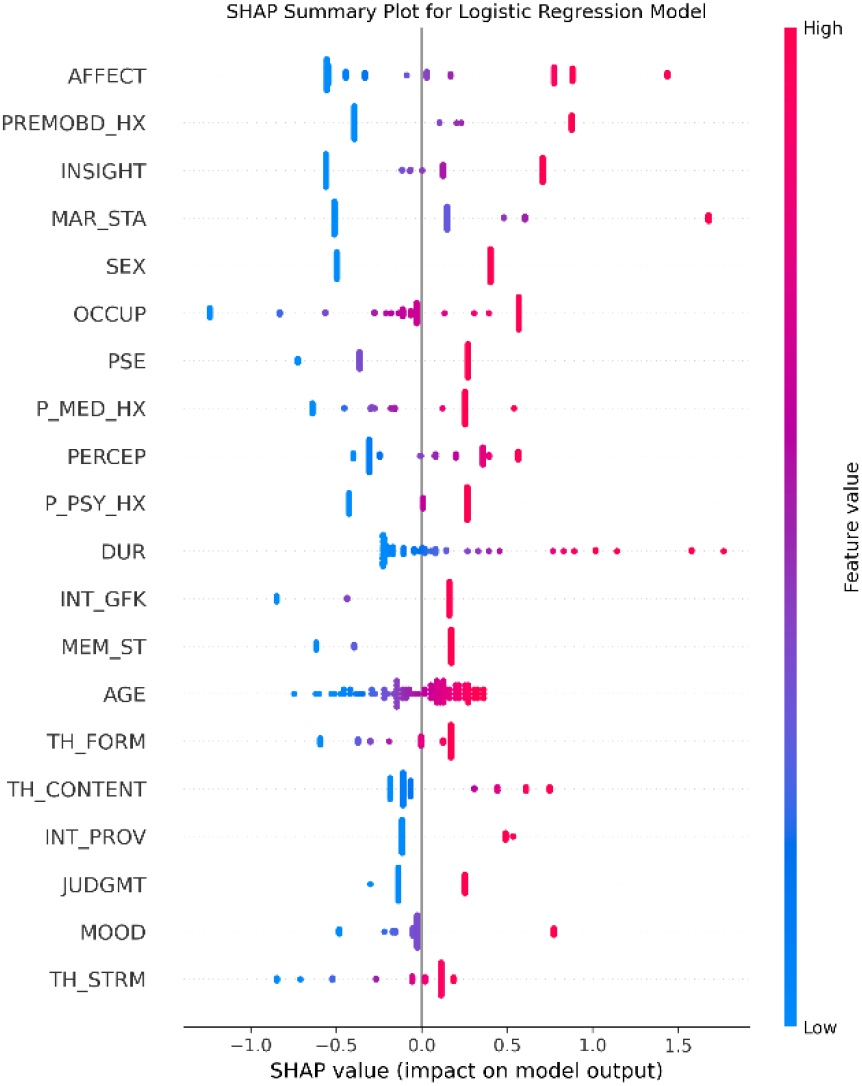
SHAP Plot for Logistic Regression model

**Figure 11.**
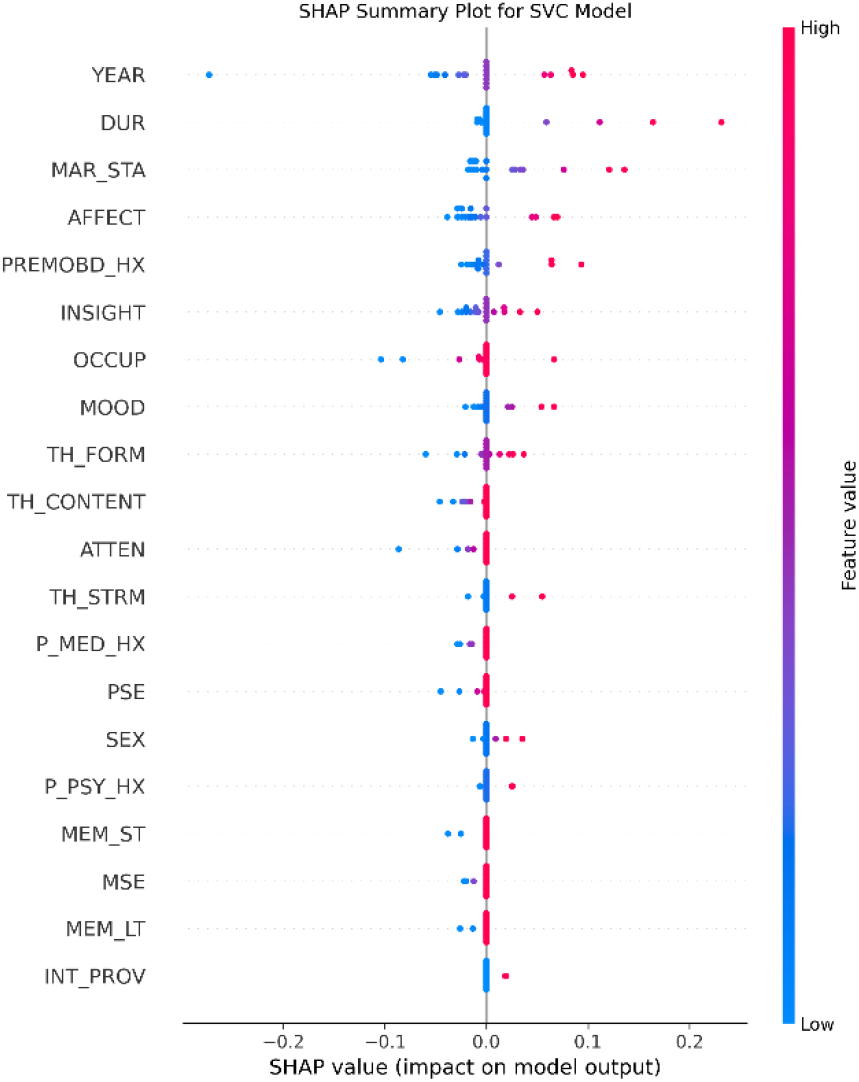
SHAP Plot for Support Vector Classification model

**Figure 12.**
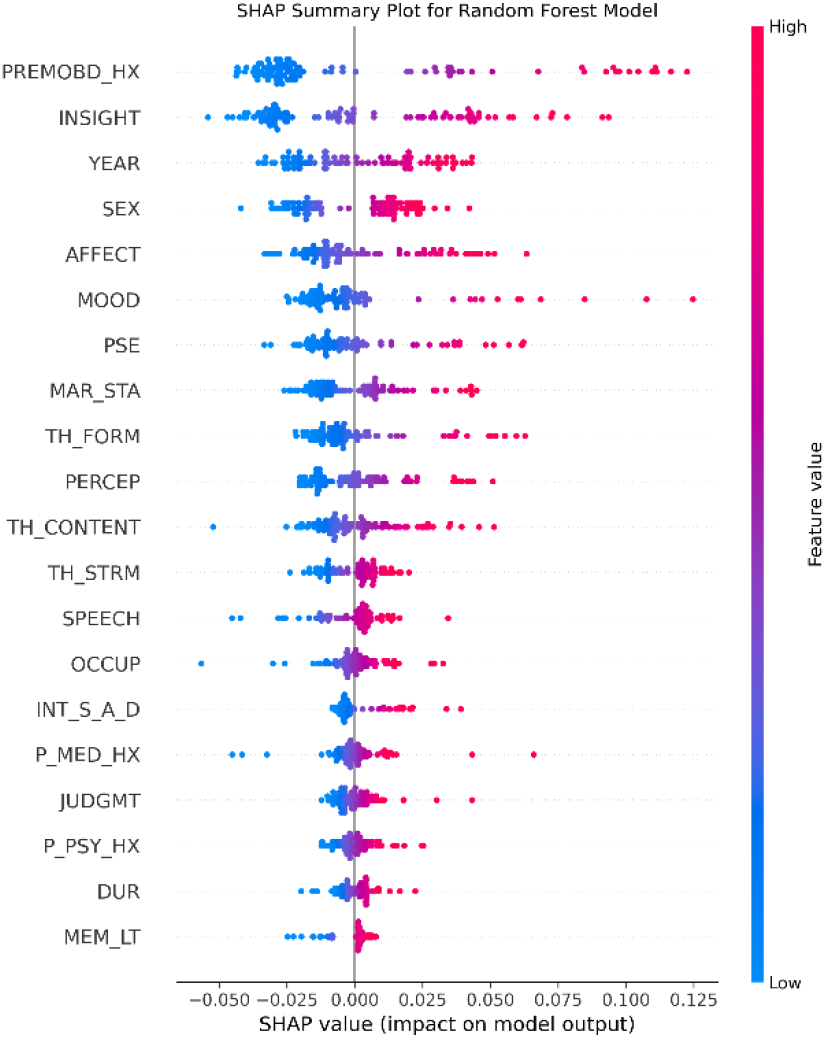
SHAP Plot for Random Forest model

**Figure 13.**
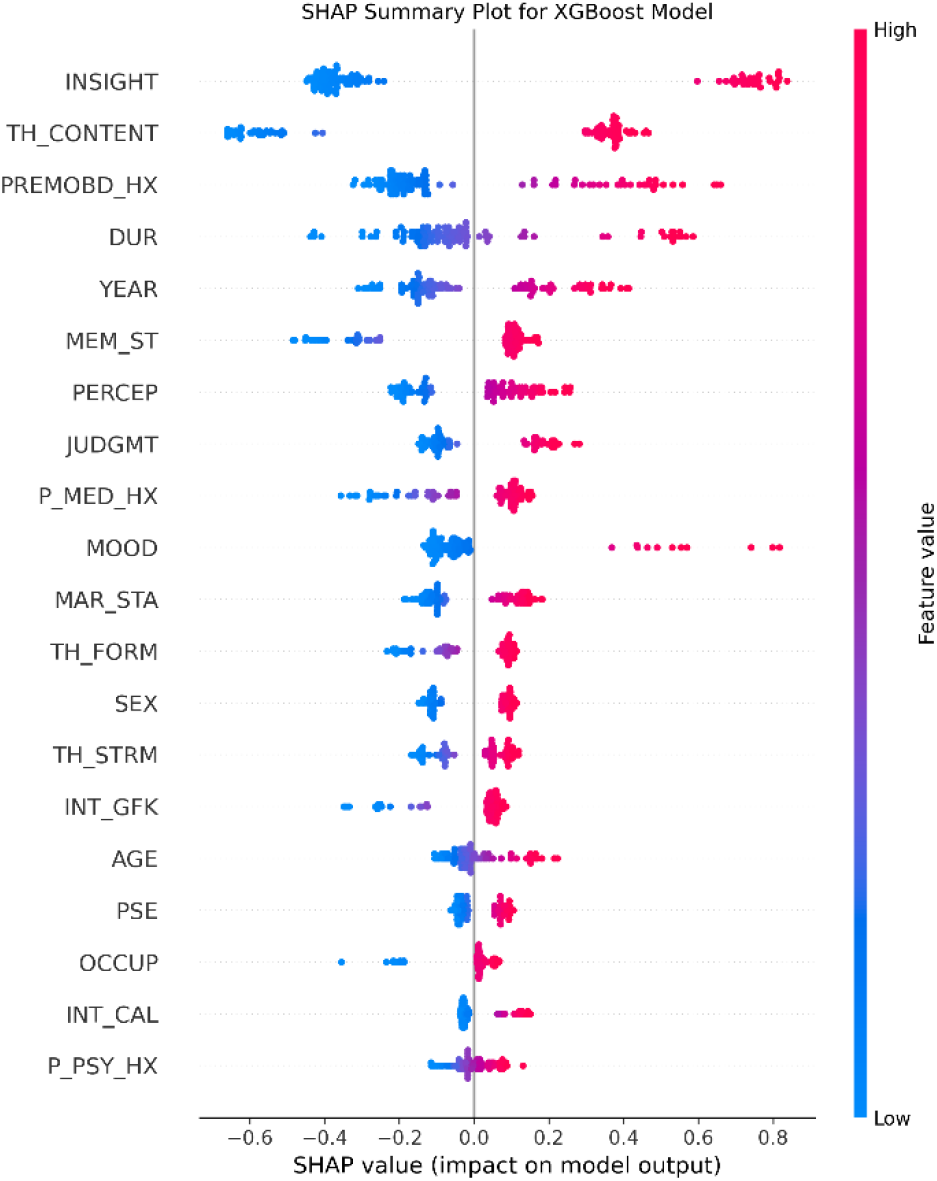
SHAP Plot for Extreme Gradient model

## 5. Conclusion

In predicting psychotic and mood disorders using a multi-model approach, the result showed that Random Forest (0.8585) performed best, followed by XGBoost model (0.8553), then Support Vector Classification (0.8217) and Logistic Regression (0.8032). The Psychotic Disorders class was well predicted (100%), while Neuro Disorders, Mood Disorders and Substance-Related Disorders were oftentimes misclassified as Psychotic Disorders. This misclassification can be attributed to class imbalance, and overlap in symptomatology. The SHAP plots revealed that Mental Status Examination, and Demographic and Functionality Groups indicators were the most influential followed by Psychiatric and Personal History. Thus, revealing that features such as insight, affection, mood, perception, occupation, marital status, year, and premorbid history were the most prominent in predicting psychotic and mood disorders.

The results demonstrate computational evidence proving that diagnostic instability in psychiatric conditions is traceable to predictable patterns. For instance, psychotic disorders showed more stable features with up to 98.6% recall, mixed-psychotic mood disorders showed less stable features with 20% recall proving that such conditions are quite algorithmically indistinguishable. Furthermore, this research provides empirical support for the spectrum hypothesis. While conventional diagnostic criteria seeks to clearly differentiate psychiatric conditions, the results shows computational evidence supporting that psychotic-mood disorders often fall into a spectrum of conditions. Lastly, the three-class approach of this research showed that psychiatric comorbidity patterns can be detected using machine learning models.

Future work will focus on larger and balanced clinical dataset to implement effective hyper-parameter tuning while also exploring the use of advanced deep learning techniques such as the Artificial Neural Network (ANN) model as it captures complex relationships between psychoses and mood disorders features.

## Data availability

The used and/or analysed datasets during this study are available from the corresponding author upon reasonable request.

## Ethical Considerations

Health Research Ethics Committee of Lagos University Teaching Hospital (LUTH) gave ethical approval for this work.

## Authorship contribution statement

Mercy Oyebode: Investigated the research area, Coding, Implementing, Analyzed results, Reviewed and summarised the literature, Wrote and edited the original draft, also managed the research activity planning and execution, Development of ideas according to the research aims. Racheal Rieninwa: Performed commentary and Revision. Samuel Okodeh: Data collection. Isioma Aniukwu: Ethics guidelines review and approval. Ephraim Nwoye: Performed critical review, Commentary, also managed the research activity planning and execution, Development of ideas according to the research aims.

## Declaration of competing interest

The authors declare that they have no known competing financial interests or personal relationships that could have appeared to influence the work reported in this paper.

## Funding

This research did not receive any specific grant from funding agencies in the public, commercial, or not-for-profit sectors.

## Notes

### Competing Interest Statement

The authors have declared no competing interest.

